# Brain cortical changes are related to inflammatory biomarkers in hospitalized SARS-CoV-2 patients with neurological symptoms

**DOI:** 10.1101/2022.02.13.22270662

**Authors:** Gretel Sanabria-Diaz, Manina Maja Etter, Lester Melie-Garcia, Johanna Lieb, Marios-Nikos Psychogios, Gregor Hutter, Cristina Granziera

**Affiliations:** Translational Imaging in Neurology (ThINK) Basel, Department of Medicine and Biomedical Engineering, University Hospital Basel and University of Basel, Basel, Switzerland; Brain Tumor Immunotherapy Lab, Department of Biomedicine, University Hospital Basel and University of Basel, Basel, Switzerland; Division of Neurosurgery, University Hospital Basel, Basel, Switzerland; Department of Neurology, University Hospital Basel, Basel, Switzerland; Department of Neuroradiology, Clinic of Radiology and Nuclear Medicine, University Hospital Basel, Basel, Switzerland

## Abstract

Increasing evidence shows that the brain is a target of SARS-CoV-2. However, the consequences of the virus on the cortical regions of hospitalized patients are currently unknown. The purpose of this study was to assess brain cortical gray matter volume (GMV), thickness (Th), and surface area (SA) characteristics in SARS-CoV-2 hospitalized patients with a wide range of neurological symptoms and their association with clinical indicators of inflammatory processes. A total of 33 patients were selected from a prospective, multicenter, cross-sectional study during the ongoing pandemic (August 2020-April 2021) at Basel University Hospital. Retrospectively biobank healthy controls with the same image protocol served as controls group. For each anatomical T1w MPRAGE image, the Th and GMV segmentation were performed with the FreeSurfer-5.0. Cortical measures were compared between groups using a linear regression model. The covariates were age, gender, age*gender, MRI magnetic field strength, and total intracranial volume/mean Th/Total SA. The association between cortical features and laboratory variables was assessed using partial correlation adjusting for the same covariates. P-values were adjusted using false discovery rate (FDR). Our findings revealed a lower cortical gray matter volume in orbitofrontal and cingulate regions in patients compared to controls. The orbitofrontal grey matter volume was negatively associated with protein levels, CSF-blood/albumin ratio and CSF EN-RAGE level. CSF EN-RAGE and CSF/Blood-albumin ratio, which are neuroinflammatory biomarkers, were associated with cortical alterations in gray matter volume and thickness in frontal, orbitofrontal, and temporal regions. Our data suggest that viral-triggered inflammation leads to increased neurotoxic damage in some cortical areas.

## Introduction

Increasing evidence shows that the severe acute respiratory syndrome coronavirus 2 (SARS-CoV-2) affects the brain (1). Following the viral infection, a peripherical immune system reaction leads to a hyperinflammatory response and chronic neuronal dysfunction (2). However, the consequences of SARS-CoV-2 in the brain of hospitalized patients, where neurological complications are observed in 80% of the cases, are currently unknown. The purpose of this study was to assess brain cortical gray matter volume (GMV), thickness (Th), and surface-area (SA) characteristics in SARS-CoV-2 hospitalized patients with a wide range of neurological symptoms compared to healthy controls, as well as their association with clinical indicators of inflammatory processes.

## Materials and Methods

A total of 33 patients with SARS-CoV-2 infection were enrolled from a prospective, multicenter, cross-sectional study during the ongoing pandemic (August 2020-April 2021) at Basel University Hospital (clinicaltrials.gov NCT04472013). All participants remained anonymous, and written consent was given by the patients or a legal representative. Inclusion criteria were age>18 years, a reverse transcriptase PCR (rRT-PCR)-positive SARS-CoV-2 infection, and a 3D high-resolution T1-weighted MRI sequence of the whole brain. Exclusion criteria were SARS-CoV-2 RT-PCR-negative testing and pregnancy. A subset of subjects (n=14 to 20) benefitted from cerebrospinal fluid (CSF) sampling. The laboratory test included leukocytes, lactate, protein levels, CSF-blood/albumin-ratio, and five cytokines (plasma-TRANCE, plasma-EN-RAGE, CSF-OPG, CSF-TRANCE, and CSF-EN-RAGE) that had been previously associated with SARS-CoV-2 infection severity (2). The cytokines analysis was performed using the Olink 96 target inflammation and neurology (https://www.olink.com/products-services/target/inflammation/) panels. The patients were stratified based on their neurological symptoms (class I: mild neurological symptoms; class II: moderate neurological symptoms; class III: severe neurological symptoms, Table 1).

**Table 1.** Primary demographics and clinical variables for the SARS-CoV-2 and Control group

|  | **Patients** | **Controls** | **P Value** |
| --- | --- | --- | --- |
| Numbers of subjects | 33 | 33 | na |
| Age | mean=50.45, sd= 19.13 | mean=51.64, sd=23.25 | .78 |
| Sex, Male/Female (M/F) | 13/20 | 12/21 | .80 |
| Neuro-COVID class | I (n=15), II (n=8), III (n=5) | na | na |
| CSF Leukocytes (mmol/l) | n=16 (mean=4.38, sd=3.83) | na | na |
| CSF Lactate (mmol/l) | n=15 (mean=1.89, sd=0.52) | na | na |
| CSF Protein total (mmol/l) | n=16 (mean=345.41, sd=186.37) | na | na |
| CSF Blood Albumin Ratio | n=14 (mean=6.50, sd=4.61) | na | na |
| CSF Glucose (mmol/l) | n=15 (mean=4.54, sd=1.33) | na | na |
| Plasma TRANCE | n=20 (mean=2.74, sd=0.93) | na | na |
| Plasma EN-RAGE | n=20 (mean=3.30, sd=1.37) | na | na |
| CSF OPG | n=18 (mean=9.55, sd=0.68) | na | na |
| CSF TRANCE | n=18 (mean=-0.31, sd=0.29) | na | na |
| CSF EN-RAGE | n=18 (mean=0.43, sd=0.74) | na | na |
| Weight (kg) | mean=69.06, sd=11.62 | mean=72.79, sd=15.85 | .44 |
| Height (meters) | mean=1.70, sd=0.10 | mean=1.69, sd=0.10 | .73 |
| MRI magnetic Field strength | 1.5 T (n=29)<br>3 T (n=4) | 1.5 T (n=11)<br>3 T (n=22) | na |
| TIV (cm<sup>3</sup>) | mean=1520.03, sd=179.37 | mean=1523.14, sd=146.27 | .86 |
| Mean cortical thickness (mm<sup>3</sup>) | mean=2.34, sd=0.15 | mean=2.39, sd=0.16 | .27 |
| Surface Area (mm<sup>2</sup>) | mean=1596.4, sd=198.25 | mean=1655.79, sd=163.27 | .10 |

Note.— Non-parametric Mann-Whitney U-test was applied whenever a variable for each group was not normally distributed (Shapiro-Wilk $P < 0.05$ ), or there was no homogeneity of variances (Levene's test); chi-square test for sex. Mean and standard deviation are shown for all variables except for sex and MRI magnetic Field strength. n=sample size, sd= standard deviation, CSF= Cerebrospinal fluid, TIV= Total Intracranial Volume, TRANCE= tumour necrosis factor-related activation-induced cytokine, EN-RAGE= receptor for advanced glycation end-products binding protein, OPG= osteoprotegerin, na= not applicable, cm=centimeters, mm=millimeters, kg=kilograms, mmol/l=millimole per liters, M=male, F=female, I=NeuroCOVID class 1, II=NeuroCOVID class 2, III=NeuroCOVID class 3.

Retrospectively biobank, age- and sex-matched healthy controls with the same image protocol served as controls and were obtained from the Neurology Department, University of Basel. Patient and control characteristics are presented in Table 1.

Anatomical T1wMPRAGE MRI sequences were acquired using two MRI scanners: Scanner-1: 1.5Tesla Siemens-Avanto-Fit and Scanner-2: 3Tesla Siemens-Skyra: Scanner-1: 160 slices in sagittal orientation; in-plane FOV= 256×256mm2, matrix size 256×256, voxel size of 1×1×1mm^3^. The echo (TE), repetition (TR), and inversion (TI) times were TE/TR/TI=2.8ms/2400ms/900ms, flip-angle FA=8°. Scanner-2: 160 slices in sagittal orientation; in-plane FOV= 256×240mm2, matrix size=256×240, voxel-size=1×1×1mm^3^, TE/TR/TI=2.98ms/2300ms/900ms, FA=9°.

Cortical thickness (Th) and volumetric segmentation were performed with the FreeSurfer-6.0 image analysis suite (http://surfer.nmr.mgh.harvard.edu/). In this study, we selected the Desikan-Killiany atlas (3) (33 cortical regions per hemisphere) for which the GMV(cm^3^), SA(mm^2^), and Th(mm) were assessed.

Clinical and demographic variables were compared between groups with independent t-tests, Mann-Whitney, or Chi-square tests as appropriate. GMV, SA, and Th were compared between groups using a linear regression model. Age, gender, age*gender, MRI-magnetic field-strength, total intracranial-volume (TIV)/mean-Th/Total-SA were used as covariates. The associations between cortical measures and laboratory variables were assessed using partial-correlation and the same covariates. For all analysis P-values were adjusted for multiple comparisons using false discovery rate (FDR). The statistical analysis was performed using the JASP-software (https://jasp-stats.org/) and MATLAB-software for the partial-correlation analysis (‘partialcorri.m’) (https://www.mathworks.com/).

## Results

Patients clinical characteristics and inflammatory biomarkers are reported in Table 1. There were no significant differences between groups in age, gender, Total-GMV, mean-Th, and Total-SA (all *P*>.05) (Table 1).

Patients showed lower GMV in the right rostral anterior cingulate (controls-mean=1.90; patients-mean=0.38; *P*=.46), left medial orbitofrontal (controls-mean=5.08; patients-mean=0.84; *P*=.46), and left superior frontal regions (controls-mean=22.01; patients-means=3.75; *P*=.46).

There were no significant differences between groups in Th and SA after FDR correction. In a subgroup of patients (CSF-study), blood leukocytes were negative associated with GMV in right lateral orbitofrontal (r=-.83; *P*=.03) and left inferior temporal regions (r=-.84; *P*=.03); lactate with left inferior temporal (r=-.85; *P*=.02), left rostral middle frontal (r=-.79; *P*=.04), and right lateral orbitofrontal (r=-.78; *P*=.04).

The highest number of significant negative correlations were found for protein levels (18 regions), CSF/blood/albumin-ratio (15 regions), and the EN-RAGE cytokine (17 regions). These regions were anatomically localized in frontal, orbitofrontal, and temporal regions (Fig A, Fig B).

The CSF/blood-albumin ratio correlated positively with left rostral anterior cingulate (r=-.81,*P*=0.03), left lateral orbitofrontal (r=.84; *P*=.02), left pars triangularis (r=.84; *P*=.02), left postcentral (r=.79; *P*=.03) and right paracentral (r=.80; *P*=.04) regional Th. A negative association was found with left rostral anterior cingulate (r=-.81; *P*=.03) and right caudal middle frontal (r=-.79; *P*=.04). Additionally, EN-RAGE correlated positively with the left pars triangularis (r=.87; *P*=.002) (Fig 1-A, Fig 1-C).

**Fig 1.**
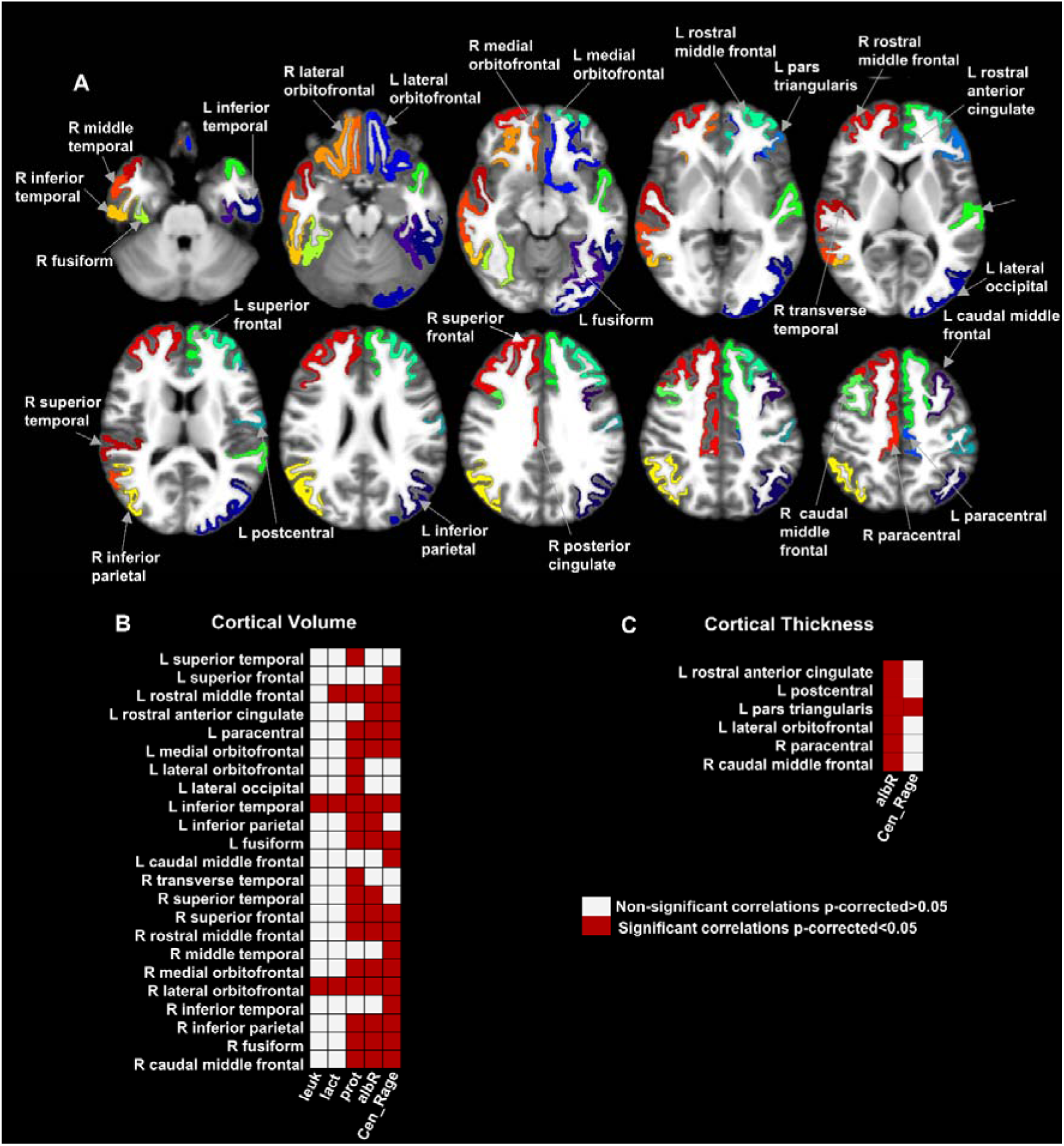
Map of the brain regions with a significant association between gray matter volume and cortical thickness clinical variables in severly affected SARS-CoV-2 patients. Panel A) show the 27 brain regions with significant correlation values for gray matter volume and cortical thickness after multiple comparison corrections (False Discovery Rate-FDR). Panel B) and C) shows the matrices representing the association significance (significant p-corrected<0.05 in red squares).CSF=Cerebrospinal fluid, leuk=leukocytes, lact=lactate, prot=protein, albR=Albumin CSF-blood ratio, Cen_Rage: CSF EN-RAGE = extracellular receptor for advanced glycation end-products binding protein, R= right, L= left.

Finally, SA showed a significant correlation between the EN-RAGE and the right posterior cingulate cortex (r=.79; *P*=.04).

## Discussion

This study shows a GMV decrease in orbitofrontal and cingulate regions in a sample of hospitalized SARS-CoV-2 patients with a wide range of neurological symptoms. These findings are in agreement and extend previous reported multifocal MRI abnormalities in hospitalized patients (4). Interestingly, the GMV in the orbitofrontal area– a region that has been described as metabolic affected in hospitalized patients (5) -was consistently negatively associated with with protein levels, CSF/blood-albumin ratio and CSF EN-RAGE.

One hypothesis for the brain alteration in SARS-CoV-2 infection is a neuroinflammatory response (6). Our results confirmed a significant association between (i) a decreased GMV and Th in frontal, fronto-orbital, and temporal regions and (ii) increased CSF levels of indirect inflammatory markers (protein, blood/albumin ratio, and EN-RAGE). Elevation in CSF protein levels has been used to indicate the inflammatory response (7). The CSF/blood-albumin ratio increase suggests a BBB breakdown. EN-RAGE is a cytokine that activates an inflammatory cascade, including accelerated atherosclerosis (8). The relationship between ENRAGE and CSF/blood-albumin ratio with increased volumes in some cortical areas suggests a viral-triggered inflammatory process as a consequence of a secondary parainfectious complication or after direct invasion (less probable) (9).

In summary, CSF inflammatory marker levels were associated with GMV and Th changes in frontal, orbitofrontal, and temporal regions in hospitalized SARS-CoV-2 patients with different levels of neurological involvement. Future longitudinal studies in larger cohorts should further confirm and expand these findings.

## Data Availability

All data produced in the present study are available upon reasonable request to the authors

## List of abbreviations

SARS-CoV-2: severe acute respiratory syndrome coronavirus 2
CSF: cerebrospinal fluid
GMV: gray matter volume
Th: cortical thickness
SA: surface area
TRANCE: tumor necrosis factor -related activation-induced cytokine
EN-RAGE: receptor for advanced glycation end-products binding protein
OPG (TNFRSF11B): osteoprotegerin
rRT-PCR: quantitative reverse transcriptase polymerase chain reaction
FDR: false discovery rate

## Acknowledgments

We would like to thank all colleagues for helping us during the current study and the selfless volunteers who participated. We are also very grateful to many members of the frontline medical staff for their selfless dedication in the face of this pandemic.

## Author contributions

Guarantors of integrity of entire study, all authors ; study concepts/study design, G.S., M.M.E., L.M., C.G.; data acquisition, J.L., M.M.E., M.P., G.H.; data analysis/interpretation, G.S., M.M.E., L.M., C.G.; manuscript drafting or manuscript revision for important intellectual content, all authors; approval of the final version of the submitted manuscript, all authors; agrees to ensure any questions related to the work are appropriately resolved, all authors; literature research, G.S., M.M.E.; clinical studies, J.L., M.M.E., M.P., G.H.; statistical analysis, G.S., L.M.; and manuscript editing, G.S., M.M.E., L.M., G.H., C.G.

* G.S. and M.M.E. contributed equally to this work.

## Disclosures of Conflicts of Interest

G.S. No relevant relationships. M.M.E. No relevant relationships. L.M. No relevant relationships. J.L. No relevant relationships. M.N.P. No relevant relationships. G.H. No relevant relationships. C.G. No relevant relationships.

## Notes

### Competing Interest Statement

The authors have declared no competing interest.

### Funding Statement

The study was funded by the BOTNAR Fast Track Call foundation grant (FTC-2020-10) awarded to G.H., Other research support to G.H. for this project include a Swiss National Science Foundation Professorial Fellowship (PP00P3_176974); the ProPatient Forschungsstiftung, University Hospital Basel (Annemarie Karrasch Award 2019); the Department of Surgery, University Hospital Basel, to G.H.
G.H. has equity in, and is a cofounder of Incephalo Inc.
CG is supported by the Swiss National Science Foundation (SNSF) grant PP00P3_176984, the Stiftung zur Forderung der gastroenterologischen und allgemeinen klinischen Forschung and the EUROSTAR E!113682 HORIZON2020.

### Author Declarations

The trial (NCT04472013) was registered under clinicaltrials.gov and approved by the local institutional review board (Ethikkommission Nordwest- und Zentralschweiz, EKNZ 2020-01503)

